# Genomic Insights into the 2023 Outbreak of Acute Hemorrhagic Conjunctivitis in Pakistan: Identification of Coxsackievirus A24 Variant through Next Generation Sequencing

**DOI:** 10.1101/2023.10.11.23296878

**Authors:** Syed Adnan Haider, Zunera Jamal, Muhammad Ammar, Rabia Hakim, Muhammad Salman, Massab Umair

**Affiliations:** National Institutes of Health, Islamabad, Pakistan

**Author notes:** **Corresponding author:** Name: Massab Umair, Address: Department of Virology, National Institutes of Health, 45500, Park Road, Chak Shahzad, Islamabad, Pakistan.

**Keywords:** Pink Eye infection, Pakistan, Outbreak, Coxsackievirus A24 variant, Acute Hemorrhagic Conjunctivitis

## Abstract

Acute Hemorrhagic Conjunctivitis (AHC) has caused a significant outbreak in Pakistan in 2023, leading to a surge in cases across various cities. To gain insights into the genetic makeup of the virus responsible for the outbreak, the National Institute of Health, Islamabad conducted a study involving twenty individuals with suspected conjunctivitis in Islamabad. Five of these samples underwent whole-genome sequencing for the first-time in Pakistan, revealing Coxsackievirus A24 variant (CVA24v), genotype IV. Phylogenetic analysis revealed a higher homology (99.38-99.43%) with isolates from Zhongshan, China, in June 2023, with notable mutations concentrated in the 3D and VP1 regions. The sample also exhibited a 94% homology with 2015 France isolates. Comparing the 2023 study sequence with partial 2005 Pakistan sequences revealed divergence in the VP1 region, marked by three non-synonymous mutations (“H25P,” “I89V,” and “I90L”), while no mutation was found in the VP3 region. Notably, no intertypic recombination events were detected. This study underscores the significance of genomic surveillance for effective understanding and management of infectious disease outbreaks, offering crucial insights for public health interventions.

## Introduction

Acute Haemorrhagic Conjunctivitis (AHC) is a highly contagious self-limiting eye condition that often leads to outbreaks. It is defined by symptoms that usually appear after an incubation period of 24 to 48 hours and remain for three to seven days. These symptoms include swollen eyelids, heavy tearing, and conjunctival hemorrhage. It is frequently connected to intimate interactions in public places like jails, schools, and swimming pools. Human enterovirus 70 (EV-70) and human coxsackievirus A24 variant (CV-A24v) are the two main enteroviruses that cause AHC [1]. Members of the Picornaviridae family’s Enterovirus genus include both EV-70 and CV-A24v [2].

CVA24v is a positive-sense, non-enveloped, single-stranded RNA genome with a length of around 7.4 kilobases. At the 5’ end of this RNA lies an untranslated region (UTR) consisting of 740 base pairs. The RNA generates a single polyprotein that is subsequently split into seven non-structural viral proteins (2A-2C, 3A-3D) and four structural capsid components (VP1-VP4).

AHC was initially documented in Ghana in 1969, with the causative agent identified as EV70 [3, 4]. A subsequent outbreak emerged in Singapore in 1970, attributed to the coxsackievirus A24 variant (CVA24v) [5, 6]. Over the past few decades, AHC has become a global concern, with over 10 million reported cases[7]. Notably, Asia and South America have witnessed several significant outbreaks. South Korea experienced its largest AHC outbreak in 2002 [8]. There was a nationwide epidemic in Thailand in 2014 [9]. By 2017, the disease had reached Mexico and rapidly spread to its Caribbean neighbors and other South American countries[10].

Recently, countries including Vietnam, India, Ghana, and Pakistan have been grappling with AHC outbreaks in 2023 [11, 12]. Pakistan has a history of confronting several such outbreaks i.e., in 1981 and in 1986, whose etilogy was linked to CVA24v [13-15] A subsequent major outbreak between 2004 to 2005, was also attributed to CVA24v. In September 2023, and Pakistan is witnessing a surge in conjunctivitis cases spanning multiple cities. To unravel the mystery behind this recent surge, our study employed Next generation sequencing (NGS) metagenomic approach using conjunctival swabs to identify the etiological agent responsible for outbreak, aiming to enhance our understanding and inform public health responses.

## Materials and methods

### Samples collection

The current study, conducted in September 2023, aimed to investigate conjunctivitis cases in patients (n=15) with symptoms such as swollen eyelids, excessive tearing, and bleeding in the conjunctiva. Multiple hospitals in the Federal Capital Islamabad participated in this study by referring their samples to the National Institute of Health. These samples were stored at -20°C until further processing. Moreover, ethical approval for the study was obtained from the Institutional Review Board of the National Institute of Health.

### Viral RNA extraction

Whole viral RNA from specimens was extracted was extracted using MagMAX™ Viral/Pathogen

Nucleic Acid Isolation Kit and KingFisher™ Flex Purification System (Thermo Scientific) according to the manufacturers’ instructions.

#### Whole-genome sequencing

In this study, a subset of 05 samples were randomly selected for shotgun metagenomic sequencing. Culturing of these samples was not performed. We then prepared it for unbiased paired-end sequencing (2 × 150 bp) using the NEBNext Ultra II Directional RNA Library Prep Kit from New England Biolabs, following the provided guidelines. To barcode the libraries, we used NEBNext Multiplex Oligos for Illumina—Dual Index Primers Set 1 from the same manufacturer. We measured the libraries’ concentration using a Qubit 4.0 fluorometer with a Qubit dsDNA HS assay kit from Invitrogen. The library size was determined with an Agilent Bioanalyzer and a DNA 1000 Kit from Agilent Technologies. We diluted each library to 2 nM concentration and combined them in equal amounts. This pool was then denatured using 0.2 N sodium hydroxide and further brought down to a 10 pM solution as per the recommended protocol. We concluded the process by sequencing the denatured libraries on the Illumina MiSeq system using the MiSeq Reagent Kit v2 (300-cycles) at the Department of Virology, NIH, Islamabad, Pakistan.

#### NGS Data Analysis

We used the FastQC v0.11.9 software [16] to evaluate the quality of the raw NGS reads we acquired. To remove low-quality bases and adapter sequences, we then employed the Trimmomatic tool (v0.39) with specific settings (ILLUMINACLIP:adapters-PE.fa:2:30:10 LEADING:3 TRAILING:3 SLIDING WINDOW:4:30 MINLEN:50) [17]. The PICARD tool (Picard MarkDuplicates) helped us eliminate PCR duplicates from these refined reads [18]. Using the SPAdes program v3.15.5, we assembled the filtered reads into contigs under standard parameters [19]. We mapped these contigs to the NCBI Non-redundant (NR) database using the Burrows-Wheeler Aligner (BWA) [20]. For calling consensus genomes, we turned to the Geneious Prime software v.2022.2, adjusting the settings to: threshold = 0%, Assign Quality = total, and minimum coverage >10 [21]. We have uploaded the consensus genome we derived to GenBank, which has assigned it the accession number OR633288.

### Phylogenetics Analysis

For phylogenetic analysis, both the entire genome sequence (around 7400 bp) and its VP1 region (915bp) obtained in this study were put through a BLAST search to identify closely related sequences. The most closely matched sequences from the BLAST results were fetched from NCBI GenBank, accessed on October 01, 2023. Our selection was based on criteria like sequence completeness, having less than 2% N’s, and the presence of the full collection date. Sequences identical to or differing by just one nucleotide from those in GenBank were omitted from the tree construction. We used the MAFFT sequence aligner [22] for multiple sequence alignment. The optimal substitution model for partial sequences (VP1) was determined to be Tamura 3-parameter: T92+I using MEGA11 [23]. This model was then used in a scaled phylogenetic analysis based on the maximum-likelihood method, with a bootstrap confidence of 1000 replicates. We utilized the IQ-TREE (v2.2.0) software [24] for the phylogenetic study. The tree derived was visualized with FigTree (v1.4.4) [25]. The entire genome sequence tree was constructed similarly, but the best-fit substitution model used was Tamura 3-parameter: T92.

Additionally, we investigated recombination events within our study sequences by examining the 5′UTR and the non-structural segment of the genome. We extracted nucleotide sequences from the 5′UTR, 2A-C, and 3A-D regions of the genome. Each of these sequences was then individually subjected to BLAST analyses to identify the most closely related sequences in GenBank for each specific region.

## Results

In September 2023, conjunctival swabs from fifteen individuals of suspected viral conjunctivitis diagnosed by physicians through clinical diagnosis were received at the NIH from various hospitals of Islamabad. Among the received samples, five samples underwent whole genome sequencing, resulting in the successful sequencing of three samples. One of the viral conjunctivitis samples exhibited 99.77% coverage when compared with reference genome OR361387.1. Additionally, the remaining two sequenced samples had 95.8% and 75.8% coverage, respectively. All the successfully sequenced samples were identified with coxsackievirus A24 variant (CVA24v) responsible for conjunctivitis. The analysis of these samples revealed that the sequenced genomes belong to genotype IV as per the Enterovirus Genotyping Tool Version 0.1 (RIVM) of coxsackievirus A24 variant (CVA24v). The sample with high coverage (99.77%) was selected for downstream analysis (NCBI Accession: OR633288).

### Phylogenetic and Mutation Analysis

The phylogenetic analysis of whole genome sample (OR633288) from the 2023 pink eye outbreak in Islamabad exhibited a higher homology of 99.38% to 99.43% at nucleotide level with recently sequenced isolates (June 2023) from Zhongshan city of China (OR361388.1, OR361387.1, OR361389.1, OR361390.1) (**Figure 1**). As compared to the China isolate (OR361388.1), majority of the nucleotide mutations in the study samples were present in 3D (n=10) and VP1 (n=9) region, followed by VP2 (n=4), 2C (n=4), 3C (n=4), 2A (n=3), 3A (n=3), VP3 (n=3) region **(Table 1)**. Additionally, the sample exhibited a nucleotide-level homology of approximately 94% with isolates from France of 2015 (KR478685.1, KR399988.1).

**Figure 1.**
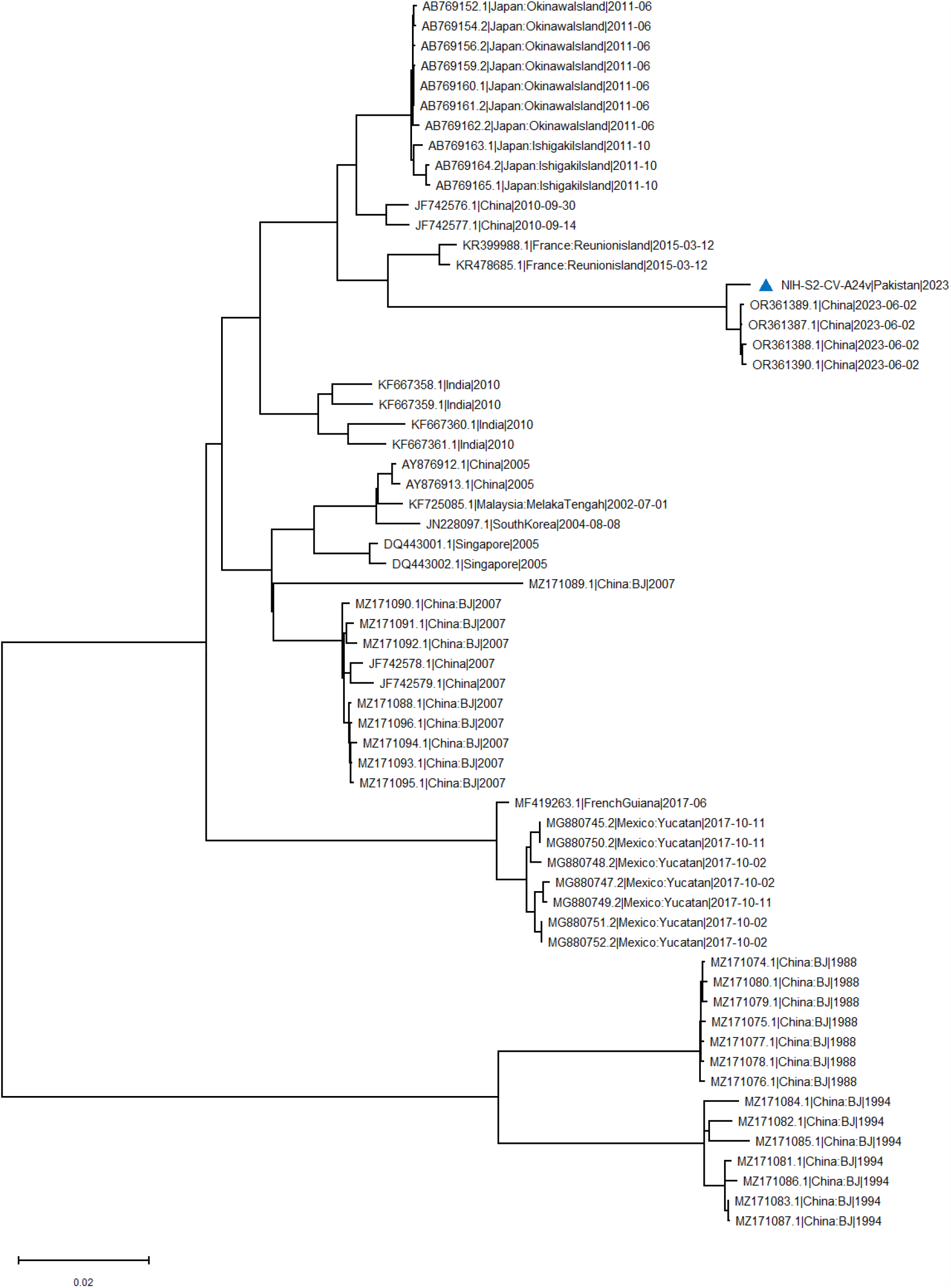
Maximum likelihood phylogenetic tree of coxsackievirus A24 variant (CVA24v) full genome sequences. The tree was generated through an IQtree with 1000 bootstraps. The study sample is marked with a blue triangle.

**Table 1.**
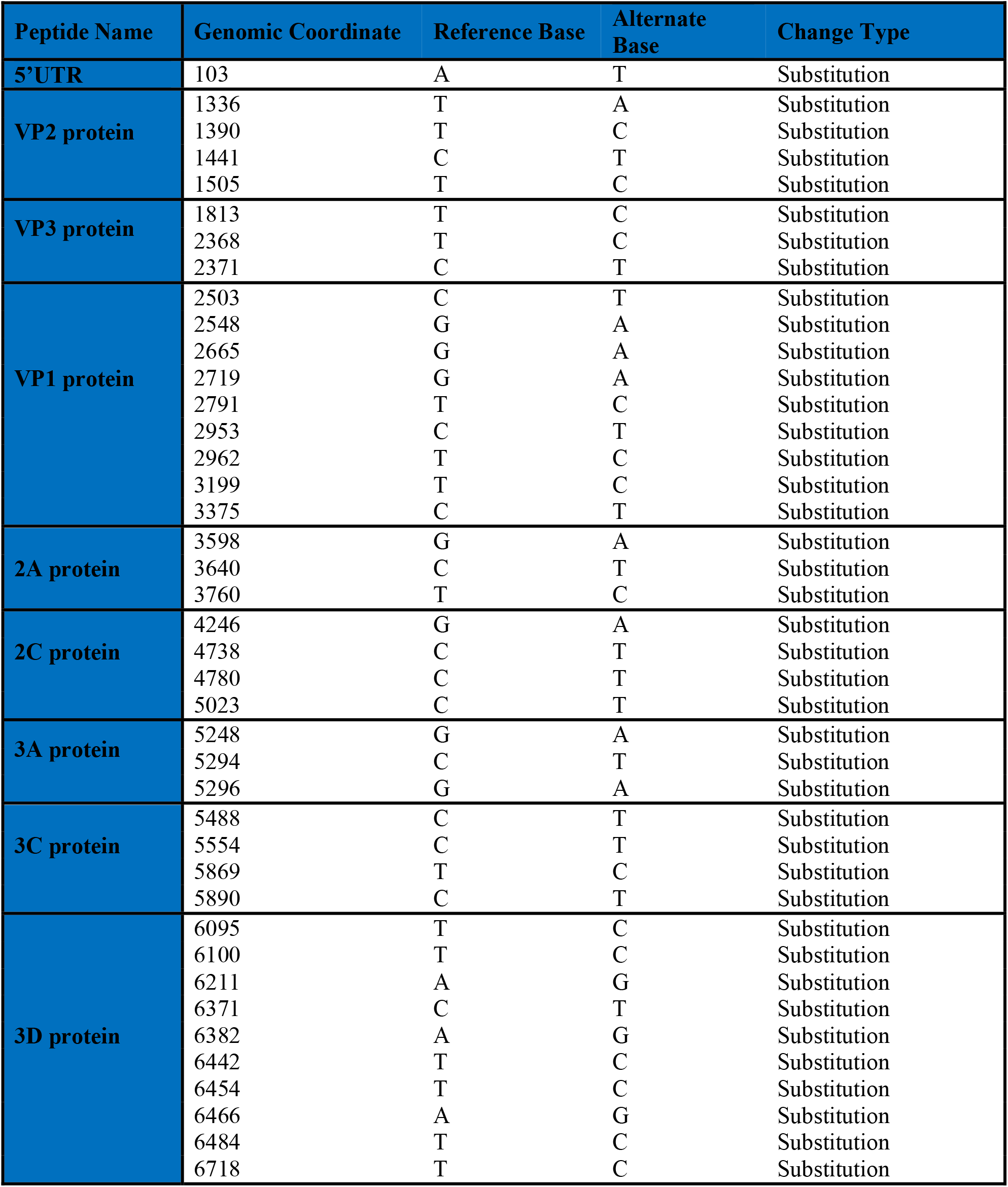
Mutation Analysis of study coxsackievirus A24 variant (CVA24v) whole genome sequence as compared to the China strain (OR361388.1). Majority of the nucleotide mutations in the study samples were present in 3D (n=10) and VP1 (n=9) region, followed by VP2 (n=4), 2C (n=4), 3C (n=4), 2A (n=3), 3A (n=3), VP3 (n=3) region

| Peptide Name | Genomic Coordinate | Reference Base | Alternate Base | Change Type |
| --- | --- | --- | --- | --- |
| **5'UTR** | 103 | A | T | Substitution |
| **VP2 protein** | 1336 | T | A | Substitution |
|  | 1390 | T | C | Substitution |
|  | 1441 | C | T | Substitution |
|  | 1505 | T | C | Substitution |
| **VP3 protein** | 1813 | T | C | Substitution |
|  | 2368 | T | C | Substitution |
|  | 2371 | C | T | Substitution |
| **VP1 protein** | 2503 | C | T | Substitution |
|  | 2548 | G | A | Substitution |
|  | 2665 | G | A | Substitution |
|  | 2719 | G | A | Substitution |
|  | 2791 | T | C | Substitution |
|  | 2953 | C | T | Substitution |
|  | 2962 | T | C | Substitution |
|  | 3199 | T | C | Substitution |
|  | 3375 | C | T | Substitution |
| **2A protein** | 3598 | G | A | Substitution |
|  | 3640 | C | T | Substitution |
|  | 3760 | T | C | Substitution |
| **2C protein** | 4246 | G | A | Substitution |
|  | 4738 | C | T | Substitution |
|  | 4780 | C | T | Substitution |
|  | 5023 | C | T | Substitution |
| **3A protein** | 5248 | G | A | Substitution |
|  | 5294 | C | T | Substitution |
|  | 5296 | G | A | Substitution |
| **3C protein** | 5488 | C | T | Substitution |
|  | 5554 | C | T | Substitution |
|  | 5869 | T | C | Substitution |
|  | 5890 | C | T | Substitution |
| **3D protein** | 6095 | T | C | Substitution |
|  | 6100 | T | C | Substitution |
|  | 6211 | A | G | Substitution |
|  | 6371 | C | T | Substitution |
|  | 6382 | A | G | Substitution |
|  | 6442 | T | C | Substitution |
|  | 6454 | T | C | Substitution |
|  | 6466 | A | G | Substitution |
|  | 6484 | T | C | Substitution |
|  | 6718 | T | C | Substitution |

The phylogenetic analysis of the VP1 region also showed higher homology (99.02%) with isolates the from China (year 2023), approximately 95% with those from Thailand (year 2014), and around 94.8% with isolates from France (year 2015) (**Figure 2**). Moreover, the VP1 region of the studied Pakistani sample from 2023 exhibited divergence from isolates dated back to 2005 in Pakistan. Notably, three non-synonymous mutations, namely “H25P,” “I89V,” and “I90L,” were identified in the VP1 region. In contrast, no mutations were detected in the VP3 region when compared to the previously available partial CDS region of Pakistani sequences from 2005. We observed no intertypic recombination events with other EVs in our analysis

**Figure 2.**
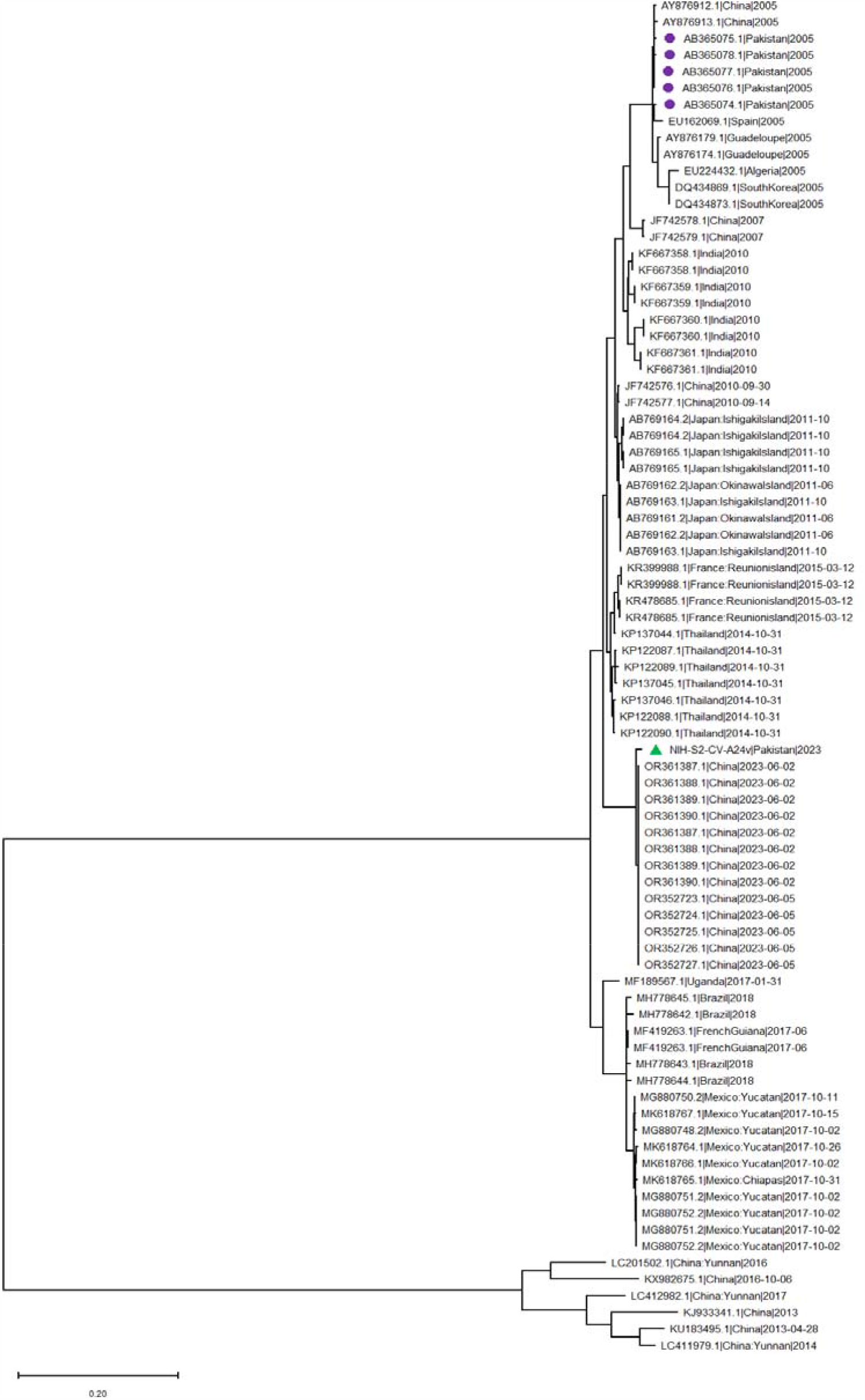
Maximum likelihood phylogenetic tree of coxsackievirus A24 variant (CVA24v) of VP1 Region. The tree was generated through an IQtree with 1000 bootstraps. The study sample is marked with a blue triangle

## Discussion

AHC has been a recurrent global health concern, with Pakistan experiencing multiple outbreaks over the years. Pakistan is experiencing an unprecedented recent surge of conjunctivitis cases, started from mid-August 2023, spreading from Karachi to various cities and provinces. By September 2023, Punjab alone reported a staggering 395,929 cases of conjunctivitis in 36 districts[26]. This massive outbreak closes 56,000 schools as 13,000 new cases were reported in a single day [27]. To know the insights of the outbreak, the current study was conducted for the first time from Pakistan that employed a Next Generation Sequencing (NGS) metagenomic approach, through which we identified CVA24v as the causative agent in all the successfully sequenced samples. This finding is consistent with Pakistan’s historical data, where outbreaks in 1986, and 2004-2005 were attributed to CVA24v through virus isolation and sequencing [14]. The recurrence of these outbreaks underscores the importance of continuous surveillance and monitoring of AHC, especially in regions with a documented history of the disease.

Our phylogenetic analysis revealed that the sequenced genomes from the 2023 outbreak in Pakistan belonged to genotype IV, which was involved in the large AHC outbreaks in South America and the Caribbean in 2017[28]. This genotype, identified since the early 2000s, has been observed to circulate consistently, contributing to various outbreaks worldwide [8, 29-31]. This underscores its ongoing prevalence and the potential to cause further outbreaks. The significant nucleotide homology of 99.38% to 99.43% observed between the Pakistani isolates and those from China collected in June 2023, implies potential epidemiological links. This also suggests a recent transmission of CVA24v between the two regions.

We further compared VP1 and VP3 region of our study Pakistani sequences of 2023 with the partial CDS region of Pakistani sequences from 2005. We found three distinct non-synonymous mutations, namely “H25P,” “I89V,” and “I90L,” in the VP1 region, known for its role in viral attachment and host cell entry. These mutations may carry implications for viral infectivity, transmission dynamics, or evasion of the immune response[32].

Given the frequent recombination events in the 5′UTR and non-structural sections of enteroviruses, we conducted a BLAST analysis on our study sequences to identify such occurrences. Our findings revealed no intertypic recombination events, consistent with the ongoing evolution of the genotype IV CV-A24v strains [33].

In conclusion, our study provides a comprehensive understanding of the recent AHC outbreak in Pakistan, emphasizing the importance of genomic surveillance in tracking and understanding the spread of infectious diseases. The close genetic relationship between the 2023 Pakistani isolates and those from other Asian regions, for instance China, France, and Thailand, calls for public health interventions to curb the spread of AHC. Future studies should focus on understanding the functional implications of the identified mutations and developing strategies for prevention and control of AHC outbreaks.

## Data Availability

Sequence generated in the current study is submitted to the GenBank NCBI are available at https://www.ncbi.nlm.nih.gov/genbank/ under the accession numbers: OR633288.

## Acknowledgement

We gratefully acknowledge the New Variant Assessment Platform (NVAP), UK Health Security Agency (UKHSA) for their support of sequencing reagents.

## Data Availability

Sequence generated in the current study is submitted to the GenBank NCBI are available at “https://www.ncbi.nlm.nih.gov/genbank/” under the accession numbers: OR633288.

## Ethics statement

The study was approved by the Internal Review Board of the National Institute of Health (NIH), Islamabad. Informed consent was obtained from all subject involved in the study

## REFERENCES

1. Wright, P.W., G. Strauss, and M. Langford, Acute hemorrhagic conjunctivitis. American family physician, 1992. 45(1): p. 173–178.

2. Treanor, J., et al., Clinical virology. 2002, ASM press.

3. Kono, R., Apollo 11 disease or acute hemorrhagic conjunctivitis: a pandemic of a new enterovirus infection of the eyes. American journal of epidemiology, 1975. 101(5): p. 383–390.

4. Mirkovic, R., et al., Enterovirus type 70: the etiologic agent of pandemic acute haemorrhagic conjunctivitis. Bulletin of the World Health Organization, 1973. 49(4): p. 341.

5. Mirkovic, R.R., et al., Enterovirus etiology of the 1970 Singapore epidemic of acute conjunctivitis. Intervirology, 1974. 4(2): p. 119–127.

6. Lim, K. and M. Yin-Murphy, An epidemic of conjunctivitis in Singapore in 1970. Singapore Med J, 1971. 12: p. 5,247.

7. Baggen, J., et al., Role of enhanced receptor engagement in the evolution of a pandemic acute hemorrhagic conjunctivitis virus. Proceedings of the National Academy of Sciences, 2018. 115(2): p. 397–402.

8. Oh, M.-d., et al., Acute hemorrhagic conjunctivitis caused by coxsackievirus A24 variant, South Korea, 2002. Emerging infectious diseases, 2003. 9(8): p. 1010.

9. Chansaenroj, J., et al., Epidemic outbreak of acute haemorrhagic conjunctivitis caused by coxsackievirus A24 in Thailand, 2014. Epidemiology & Infection, 2015. 143(14): p. 3087–3093.

10. Sousa, I.P., et al., Re-emergence of a coxsackievirus A24 variant causing acute hemorrhagic conjunctivitis in Brazil from 2017 to 2018. Archives of virology, 2019. 164: p. 1181–1185.

11. Harp, M.D. Conjunctivitis (pink eye) sees a spike in cases in Vietnam, India and Pakistan. September 6, 2023; Available from: https://www.ophthalmologytimes.com/view/conjunctivitis-pink-eye-sees-a-spike-in-cases-in-vietnam-india-and-pakistan.

12. Hemorrhagic Conjunctivitis: All you need to know about the viral ‘apollo’ eye infection. Health News of Sunday, 10 September 2023 October 05, 2023]; Available from: https://www.ghanaweb.com/GhanaHomePage/NewsArchive/Hemorrhagic-Conjunctivitis-All-you-need-to-know-about-the-viral-apollo-eye-infection-1841390.

13. Ghafoor, A. and M. Burney, Acute haemorrhagic conjunctivitis. 1987. p. 61–62.

14. Khan, A., et al., An outbreak of acute hemorrhagic conjunctivitis (AHC) caused by coxsackievirus A24 variant in Pakistan. Virus research, 2008. 137(1): p. 150–152.

15. Ishiko, H., et al., Phylogenetic analysis of a coxsackievirus A24 variant: the most recent worldwide pandemic was caused by progenies of a virus prevalent around 1981. Virology, 1992. 187(2): p. 748–759.

16. Andrews, S., FastQC: a quality control tool for high throughput sequence data. 2010, Babraham Bioinformatics, Babraham Institute, Cambridge, United Kingdom.

17. Bolger, A.M., M. Lohse, and B. Usadel, Trimmomatic: a flexible trimmer for Illumina sequence data. Bioinformatics, 2014. 30(15): p. 2114–2120.

18. Institute, B., “Picard Toolkit.” Broad Institute, GitHub Repository. Picard Toolkit, 2019.

19. Bankevich, A., et al., SPAdes: a new genome assembly algorithm and its applications to single-cell sequencing. Journal of computational biology, 2012. 19(5): p. 455–477.

20. Li, H. and R. Durbin, Fast and accurate long-read alignment with Burrows–Wheeler transform. Bioinformatics, 2010. 26(5): p. 589–595.

21. Kearse, M., et al., Geneious Basic: an integrated and extendable desktop software platform for the organization and analysis of sequence data. Bioinformatics, 2012. 28(12): p. 1647–1649.

22. Katoh, K. and D.M. Standley, MAFFT multiple sequence alignment software version 7: improvements in performance and usability. Molecular biology and evolution, 2013. 30(4): p. 772–780.

23. Tamura, K., G. Stecher, and S. Kumar, MEGA11: molecular evolutionary genetics analysis version 11. Molecular biology and evolution, 2021. 38(7): p. 3022–3027.

24. Nguyen, L.-T., et al., IQ-TREE: a fast and effective stochastic algorithm for estimating maximum-likelihood phylogenies. Molecular biology and evolution, 2015. 32(1): p. 268–274.

25. Rambaut, A., FigTree. Tree figure drawing tool. http://tree.bio.ed.ac.uk/software/figtree/, 2009.

26. Conjunctivitis cases continue to surge across Punjab. Oct 01, 2023.

27. Bowman, V. Mass outbreak of eye infections closes 56,000 schools in Pakistan. 3 October 2023 October 05, 2023].

28. Enfissi, A., et al., Coxsackievirus A24 variant associated with acute haemorrhagic conjunctivitis cases, French Guiana, 2017. Intervirology, 2018. 60(6): p. 271–275.

29. Ghazali, O., et al., An outbreak of acute haemorrhagic conjunctivitis in Melaka, Malaysia. Singapore Med J, 2003. 44(10): p. 511–6.

30. Dussart, P., et al., Outbreak of acute hemorrhagic conjunctivitis in French Guiana and West Indies caused by coxsackievirus A24 variant: phylogenetic analysis reveals Asian import. Journal of medical virology, 2005. 75(4): p. 559–565.

31. Chu, P.-Y., et al., Molecular epidemiology of coxsackie A type 24 variant in Taiwan, 2000–2007. Journal of clinical virology, 2009. 45(4): p. 285–291.

32. Yang, S.-L., et al., Annexin II binds to capsid protein VP1 of enterovirus 71 and enhances viral infectivity. Journal of virology, 2011. 85(22): p. 11809–11820.

33. Yen, Y.-C., et al., Phylodynamic characterization of an ocular-tropism coxsackievirus A24 variant. PLoS One, 2016. 11(8): p. e0160672.

